# AnterioR-Posterior VErsuS Anterior-LaTeral defibrillator pAd position in out of hospital cardiac aRresT

**DOI:** 10.64898/2026.08.24.26361176

**Authors:** Adam Colbourne, Tom Dart, Charles D. Deakin, Keith Couper, Christopher M Smith, Stewart Davies, Kate Hawley, Helen Pocock, Joshua Miller, Lauren Williams, Sophie Price, Nigel Rees

## Abstract

**Background:** Early defibrillation is a key factor of survival following out-of-hospital cardiac arrest (OHCA). In the United Kingdom, initial anterior–lateral (AL) defibrillator pad placement is standard practice. However, anterior–posterior (AP) pad positioning has been proposed as a method of improving current flow through the myocardium particularly in cases of refractory ventricular fibrillation (VF). Concerns remain regarding potential delays to defibrillation associated with alternative pad placement strategies and the practical application of AP positioning.

**Objective:** We conducted two consecutive simulation studies to explore whether there is a difference in the time taken to apply defibrillator pads, and placement accuracy, between AP and AL positions during simulated OHCA.

**Methods:** This research study comprised of two simulation studies. First, we evaluated pad placement accuracy before and after written instruction showing optimal pad placement (RESTART-SIM accuracy). Second, we undertook a randomised crossover examining time to pad placement, timing to successful pad application was recorded for each attempt (RESTART-SIM speed).

**Results:** RESTART-SIM (accuracy) out of 14 participants 50% correctly placed AL pads and 14% correctly placed AP pads initially. Following provision of guidance this increased to 93% for AL but remained at 14% for AP placement.

9 participants completed RESTART-SIM (speed). Mean AP pad placement time was 14.2 seconds and standard deviation of 2.64, compared with 10.6 seconds for AL placement and a standard deviation of 4.24. AP first strategy mean placement time was 15.3 seconds and mean AL placement time was 11.5 seconds. In AL first mean AP placement time was 12.8 seconds and mean AL placement time was 9.6 seconds.

**Conclusion:** AP pad placement whilst slower than AL placement the time difference of 4 seconds is unlikely to be clinically significant. However, without guidance, both AL and AP placements were often inaccurate. After a guidance picture AL placement was increased but AP remained poorly placed.

## Introduction

Out-of-hospital cardiac arrest (OHCA) is defined as a sudden cessation of cardiac mechanical activity resulting in circulatory collapse and absence of signs of life (Myat et al., 2018). OHCA remains a leading cause of mortality in the United Kingdom, with approximately 43,000 resuscitation attempts undertaken annually (Smith et al., 2025). despite significant resource, investment and public interest survival to 30 days remains low, 9.5% in England, 9.6% in Scotland and 6.5% in Northen Ireland (Smith et al., 2025)

Early recognition of OHCA, prompt initiation of cardiopulmonary resuscitation (CPR), and rapid delivery of defibrillation in shockable rhythms are well-established predictors of return of spontaneous circulation (ROSC) and favourable neurological outcomes (Olasveengen et al., 2021). As such, minimising time to defibrillation remains a central focus of OHCA management.

Anterior–lateral (AL) defibrillator pad placement is currently recommended as standard practice in European Resuscitation Council (ERC) guidelines (Soar et al., 2025). This pad positioning aims to optimise the electrical vector across the myocardium, facilitating effective energy delivery. However, evidence demonstrates that pad placement inaccuracies are common in both clinical and simulated settings (Ristagno et al., 2025). Suboptimal positioning of defibrillator pads may reduce current flow through the myocardium and negatively influence defibrillation success rates (Dennie Wulterkens et al., 2025). This has led to renewed interest in whether alternative pad configurations could improve defibrillation efficacy.

Anterior–posterior (AP) pad placement has been proposed as a potential means to defibrillate refractory ventricular fibrillation, since it offers a method of improving current flow through the myocardium. Observational research by Lupton et al. (2024) demonstrated that AP pad placement was associated with 2.64-fold greater adjusted odds of ROSC, although no statistically significant differences were observed in survival to hospital admission or discharge. Similarly, Rock et al. (2026) suggests initial non-shockable rhythms had a higher cumulative incidence of ROSC but high-quality evidence guiding optimal pad placement during cardiac arrest remains limited.

The 2025 European Resuscitation Council guidelines (2025) recommend considering AP pad placement in refractory VF after three initial shocks using the AL pad position, but important practical questions remain about AP use in the prehospital environment. A key concern is whether AP pad placement can be achieved without delaying defibrillation which may result in unacceptable interruptions and delay in CPR for those in non-shockable rhythms who may never stand to benefit from alternative pad position. Unlike AL placement, AP positioning requires moving or rolling the patient to apply the posterior pad, which could be challenging for solo responders or in physically constrained prehospital settings. Any delay to shock delivery could offset potential physiological benefits of AP positioning.

RESTART-SIM was therefore designed as pre-protocol simulation to inform development of a clinical protocol. It comprised two linked simulation studies, RESTART-SIM (accuracy) assessed the anatomical accuracy of AL and AP pad placement under unguided and written-guidance conditions. RESTART-SIM (speed) was a randomised crossover simulation study comparing the time required to apply pads in AL and AP positions. Together, these studies aimed to determine whether AP placement is both feasible and practical, and whether it introduces a clinically relevant delay compared with standard AL positioning.

### Ethics

This was a simulation study involving EMS staff only and was registered as a service evaluation with the Welsh Ambulance Service University Trust through the research and innovation department. We developed a simulation protocol which was reviewed by two of the authors (KC and CS). Participants read a participant information sheet which included a debrief and thank you for taking part and were consented into taking part.

## Methods

### Study design and reporting

RESTART-SIM (accuracy) evaluated the anatomical accuracy of anterior–lateral (AL) and anterior–posterior (AP) defibrillator pad placement under unguided and written-guidance conditions. RESTART-SIM (speed) was a separate randomised crossover simulation study comparing the time required to apply defibrillator pads in AL and AP positions during simulated out-of-hospital cardiac arrest.

The reporting of RESTART-SIM was guided by the STROBE statement for observational research and by relevant Healthcare Simulation Research reporting extensions to STROBE and CONSORT (Cheng et al., 2016). Reflecting the observational design of RESTART-SIM (accuracy) and the randomised crossover design of RESTART-SIM (speed).

## RESTART-SIM (accuracy)

### Participants

Participants were recruited as a convenience sample from suitable clinical education environments and related events across an ambulance service covering a population of 3 million people in 2025. Participants were eligible if they had prior training in basic life support and a fundamental understanding of defibrillator or automated external defibrillator use. The cohort included healthcare professionals and ambulance service staff. No exclusion criteria were applied on the basis of professional role or seniority.

All participants received a participant information sheet and verbal explanation before consenting and taking part. After the simulation there was time to discuss the correct pad positioning if the participant was still unclear and to ask any questions.

### Study Design

RESTART-SIM (accuracy) was a simulation-based observational study designed to evaluate the accuracy of defibrillator pad placement in AL and AP configurations. Performance was assessed under two conditions: unguided pad placement and pad placement following brief written instruction.

Simulations were conducted in private areas to minimise observation bias. A maximum attempt time of 60 seconds was imposed to reflect the clinical imperative for rapid defibrillation.

### Procedure

Participants were allocated an anonymised numerical identifier with variations (A–D) to obscure identity during independent review of pad placement accuracy and to clearly label guided vs unguided in each pad configuration attempt. Participants were initially asked to apply defibrillator pads in the AL position, followed by AP placement, without guidance. Each attempt was capped at 60 seconds. Pad positioning accuracy and time to placement were recorded, initially planned to be contained within one study but due to recommendation to randomise participants a second one was conducted.

Participants were then provided with a written instructional leaflet showing correct pad placement and given three minutes to review it. The placement process was then repeated under guided conditions for both AL and AP positions.

An 80-kg manikin was used throughout the study, the manikin was not clothed. To remove potential confounding factors for pad placement. Simulations were conducted in private areas and out of sight of other potential participants.

### Outcome measure

The primary outcome was anatomical accuracy of pad placement. Photographs of pad placements were reviewed independently by two assessors who were blinded to participant identity, timing data and participant characteristics. Pad placement was classified as acceptable or unacceptable, with qualitative descriptions recorded for the nature of any placement error. Timing data were also recorded, including incomplete attempts within the 60-second limit.

## RESTART-SIM (speed)

### Participants

Participants in RESTART-SIM (speed) were EMS personnel selected because of their relevant clinical experience, structured defibrillation training and familiarity with monitor-defibrillators and standard ambulance service defibrillation pads. The study was conducted during a scheduled EMS event. Participants were informed in advance, received study information, provided informed consent and did not receive payment.

#### Randomisation and procedure

RESTART-SIM (speed) used a randomised crossover design so that each participant acted as their own control. Participants were randomised using a 505/50 random generator into one of two groups: Group A completed AP pad placement followed by AL placement, while Group B completed AL pad placement followed by AP placement.

All participants received a standardised demonstration of both pad positions before testing. Each simulation began with a verbal start command. Timing started at this command and ended once both defibrillator pads were fully adhered to the manikin in the assigned configuration.

A full-body 80-kg manikin dressed in a zipped shirt and tracksuit trousers was used to improve ecological validity. Clothing was repositioned between attempts and participants. The study was conducted in a lecture-room setting, with other participants present and able to observe.

### Outcome Measure

The primary outcome measure was time to successful pad placement, defined as the elapsed time from the verbal start command to full adhesion of both defibrillator pads recorded using a stopwatch.

### Statistical analysis

Accuracy outcomes were summarised descriptively as counts and proportions of acceptable and unacceptable placements for each pad position and guidance condition. Qualitative placement errors were grouped according to anatomical error type, including inferior or abdominal placement, laterality error, midline straddling, excessive superior placement and lateral positioning error.

Timing outcomes were summarised using mean pad placement times for AP and AL configurations. Paired comparisons were used for within-participant AP versus AL timing data. Given the exploratory nature and small sample size, all statistical analyses were interpreted cautiously and considered hypothesis-generating rather than definitive.

## Results

### SIM (accuracy)

Fourteen participants completed RESTART-SIM (accuracy) between the ages of 21 and 56. Pad placement accuracy varied by pad configuration and by whether participants received written guidance.

During unguided attempts, acceptable pad placement was achieved in 7/14 (50%) anterior–lateral (AL) attempts and 2/14 (14%) anterior–posterior (AP) attempts. Following written guidance, acceptable AL placement increased to 13/14 (93%) attempts. In contrast, acceptable AP placement remained unchanged at 2/14 (14%) attempts.

Errors were common across both pad configurations, but the pattern of error differed. In unguided AL placement, errors were most frequently related to superior positioning, including overlap with the clavicle, and lateral positioning. After guidance, these errors were largely reduced. AP placement errors were more persistent. Although guidance appeared to reduce midline straddling, inferior or abdominal placement remained common and was the dominant error in guided AP attempts.

**Table 1.** RESTART-SIM (accuracy) Independently reviewed placement.

|  | AL Unguided (A) | AP Unguided (B) | AL Guided (C) | AP Guided (D) |
| --- | --- | --- | --- | --- |
| Acceptable | 7 | 2 | 13 | 2 |
| Unacceptable | 7 | 12 | 1 | 12 |
| Total | 14 | 14 | 14 | 14 |

Blue indicates optimal pad placement (not anatomically) and orange indicates unacceptable pad placement and which direction the pads was placed incorrectly.

**Figure 1.**
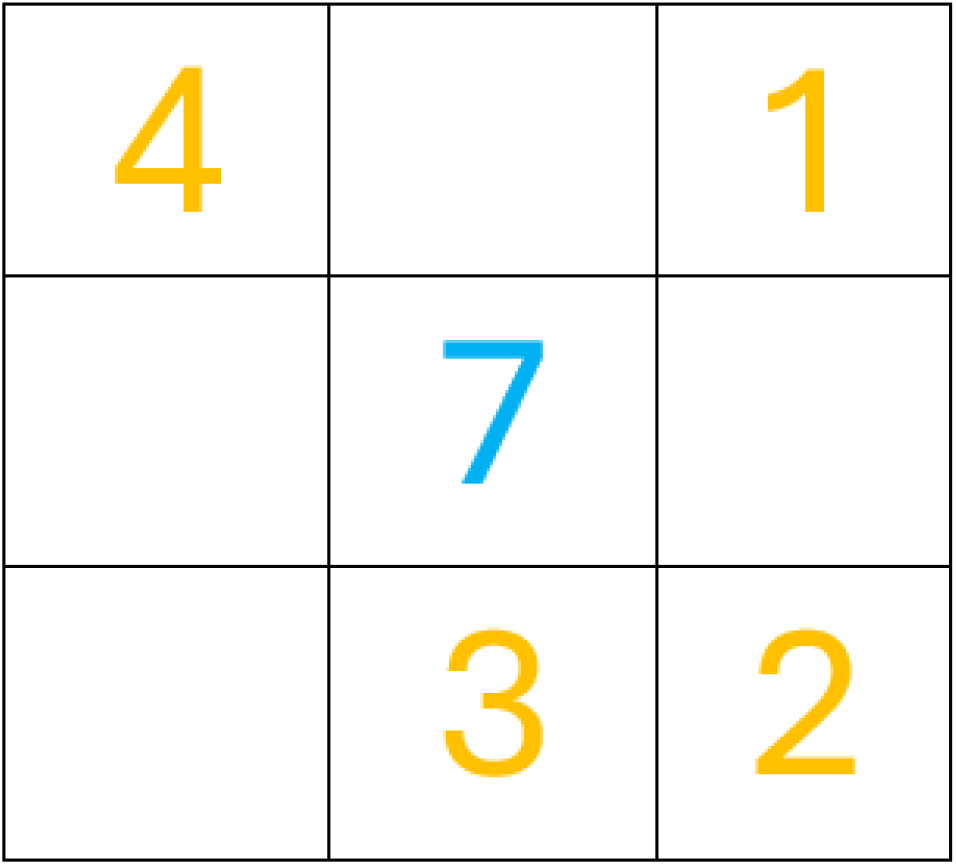
Unguided AL.

**Figure 2.**
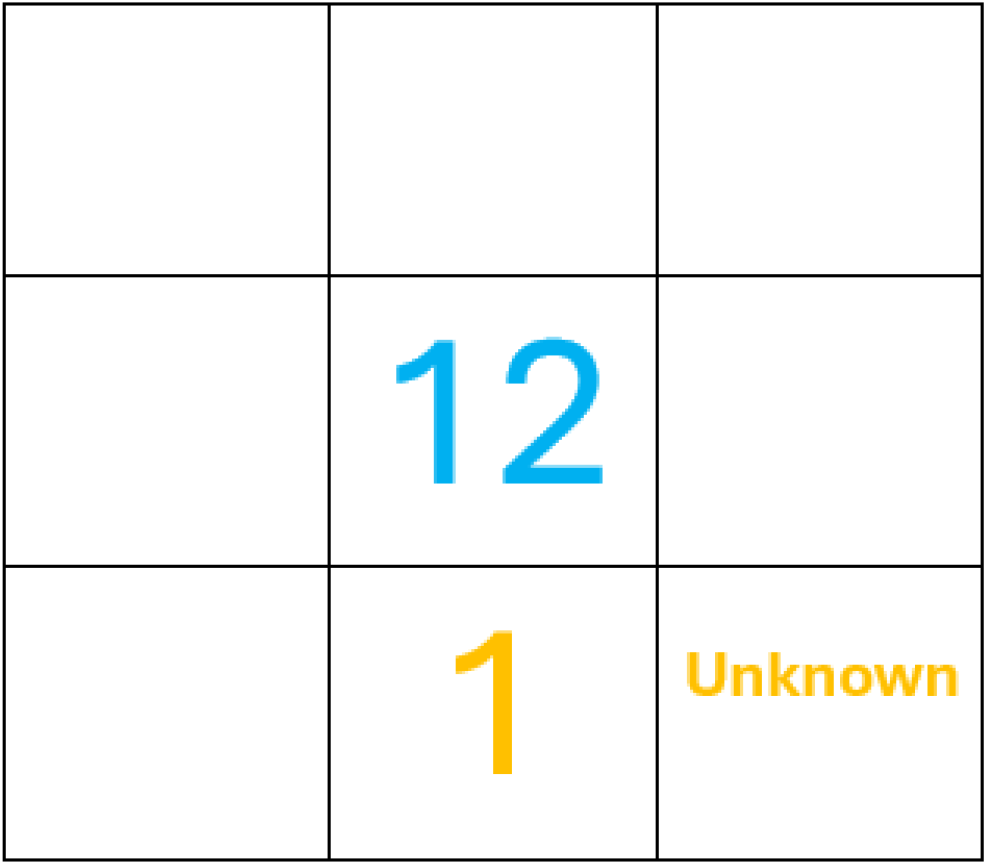
Guided AL.

**Figure 3.**
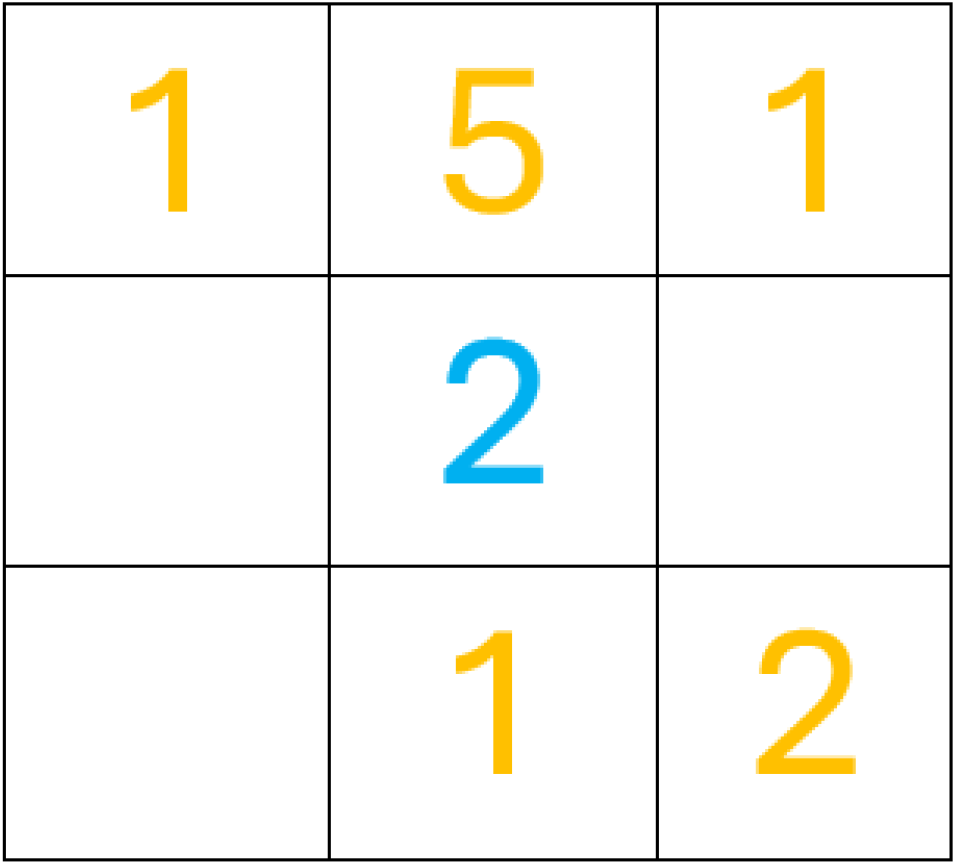
Unguided AP.

**Figure 4.**
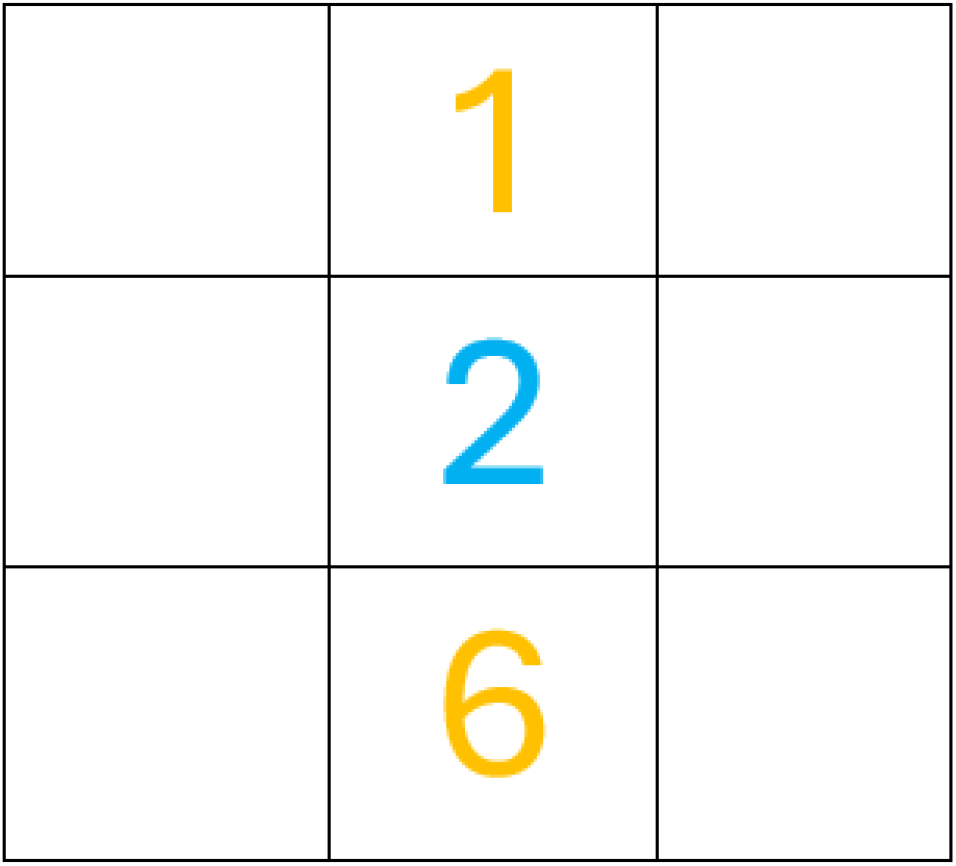
Unguided AP.

**Figure 5.**
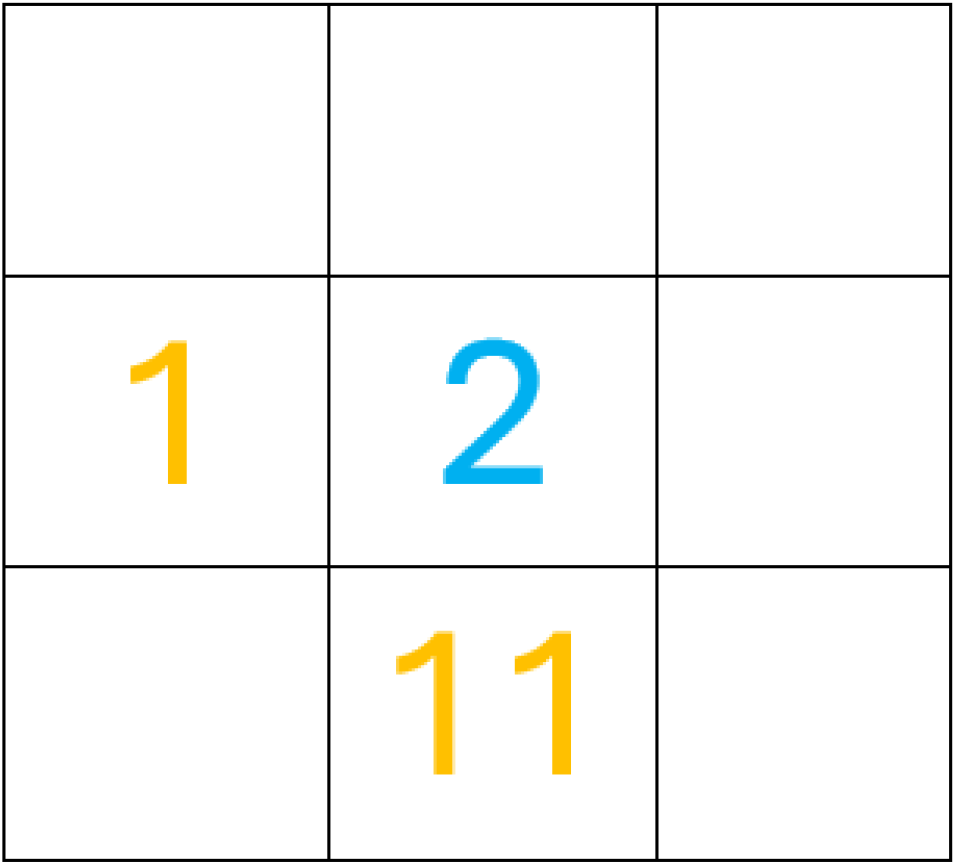
Guided AP.

**Figure 6.**
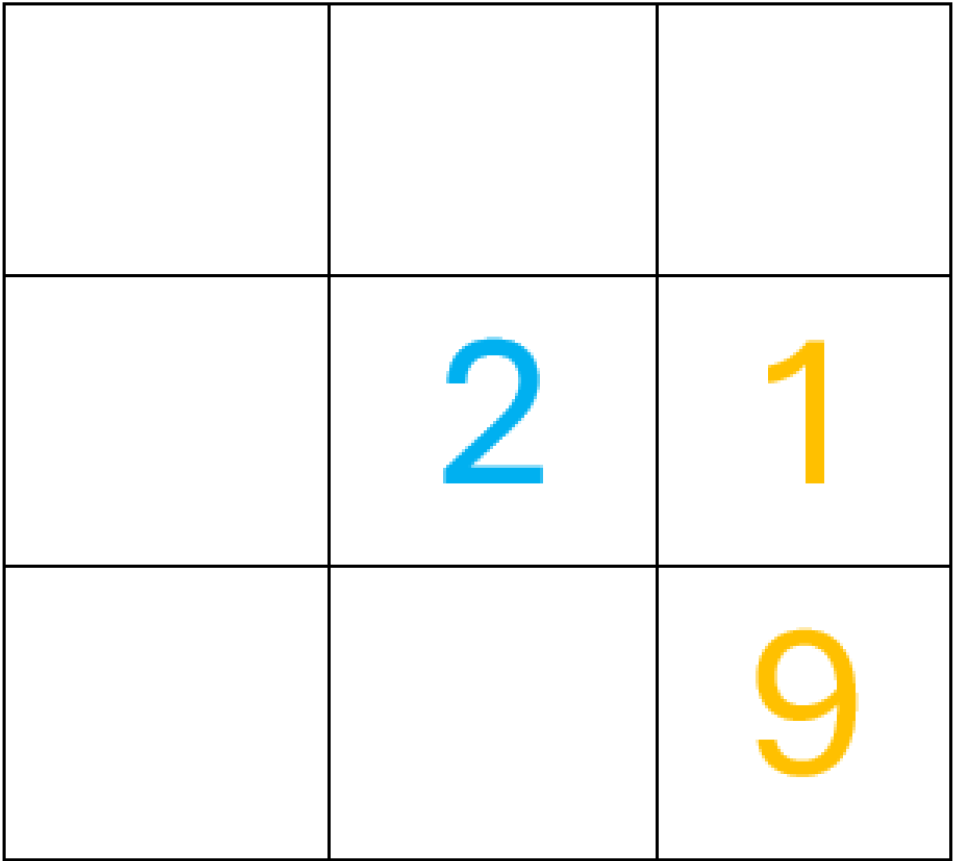
Guided AP.

### SIM (speed)

Nine EMS participants completed RESTART-SIM (speed). Five participants were randomised to Group A, completing AP placement followed by AL placement, and four participants were randomised to Group B, completing AL placement followed by AP placement.

Mean AP pad placement time was 14.19 seconds, compared with 10.62 seconds for AL placement. AP placement therefore took a mean of 3.57 seconds longer than AL placement. The paired comparison suggested this difference was statistically significant, with a p value of 0.0419.

**Table 2.**
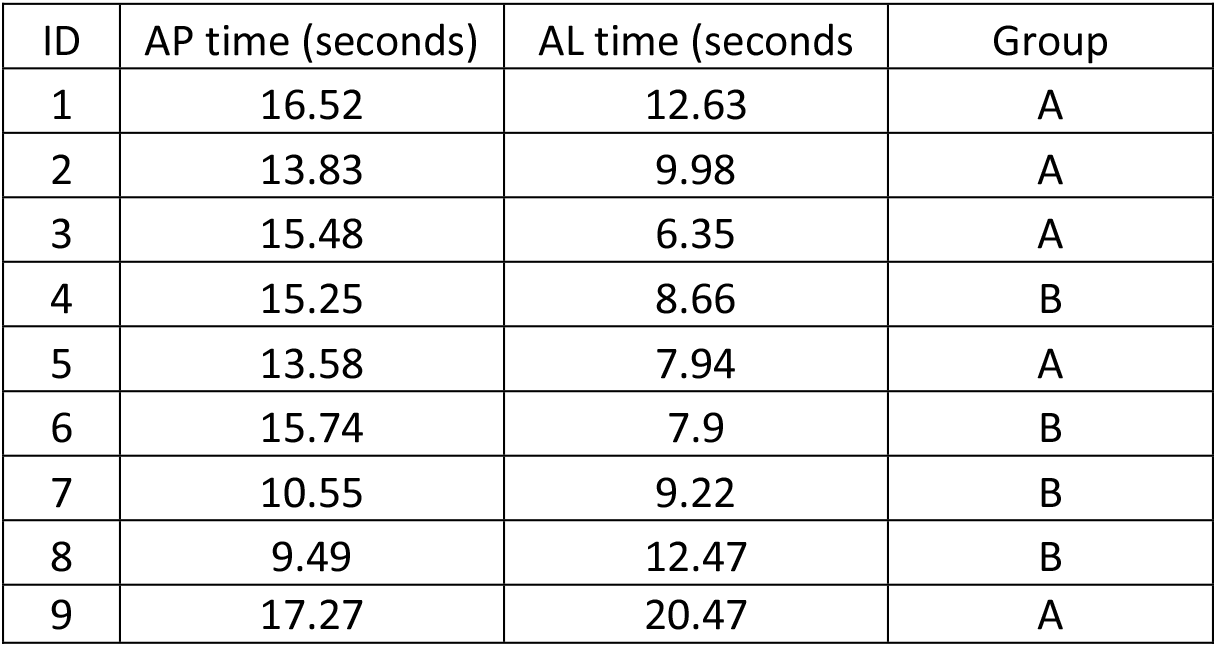
RESTART-SIM (speed) Results.

When analysed by randomisation order, both AP and AL placement times appeared longer in AP-first strategy than AL-first strategy. In Group A, mean AP placement time was 15.34 seconds and mean AL placement time was 11.47 seconds. In Group B, mean AP placement time was 12.76 seconds and mean AL placement time was 9.56 seconds.

**Table 3.** RESTART-SIM (speed) Mean time between groups.

| Group | Mean AP time (seconds) | Mean AL time (Seconds) |
| --- | --- | --- |
| A | 15.34 | 11.47 |
| B | 12.76 | 9.56 |

The current studies should therefore be interpreted as feasibility studies capable of identifying large effects and operational issues, but underpowered to exclude smaller or more moderate differences between AP and AL pad placement.

## Discussion

The primary objective of the RESTART-SIM (speed) was to determine whether AP pad placement results in a statistically significant delay when compared with standard AL positioning. This outcome is clinically significant, as any delay to defibrillation may negate potential physiological benefits associated with alternative pad placement.

Early defibrillation remains one of the strongest predictors of ROSC and favourable neurological outcomes following OHCA (Lee et al., 2021). Demonstrating that AP pad placement can be performed without introducing clinically meaningful delays is therefore a prerequisite to any significant introduction into practice.

By focusing on pad placement speed in a controlled simulation environment, the RESTART study seeks to inform future clinical trials and contribute to evidence-based guideline development while maintaining patient safety as the central priority.

The linked RESTART-SIM studies suggest that anterior–posterior (AP) defibrillator pad placement is feasible in a simulated out-of-hospital cardiac arrest setting, but may be slower and less accurately performed than standard anterior–lateral (AL) placement. This is clinically important because early defibrillation remains a key aspect of return of spontaneous circulation and favourable neurological outcome after shockable cardiac arrest (Olasveengen et al., 2021; Lee et al., 2021). Any potential physiological advantage of AP placement must therefore be balanced against the risk of delaying shock delivery.

RESTART-SIM (accuracy) showed pad misplacement was common, particularly during unguided attempts. Written guidance appeared to improve AL placement substantially, which is consistent with previous evidence that instructional diagrams can influence AED pad positioning accuracy (Foster and Deakin, 2019). However, the same improvement was not seen with AP placement. Although guidance reduced some errors, such as midline straddling, inferior or abdominal placement remained common. This suggests that AP placement may be more difficult to conceptualise anatomically and that written guidance alone may be insufficient.

These findings are consistent with wider evidence that defibrillator pad placement inaccuracies are common in clinical and simulated settings (Dennie Wulterkens et al., 2025; Missel et al., 2025). Persistent inferior AP placement is important because incorrect vertical positioning may alter the electrical vector across the myocardium and reduce effective current delivery. Future training may therefore need to emphasise anatomical landmarks more clearly, particularly for posterior pad positioning, and may require video-based instruction or supervised practical rehearsal.

RESTART-SIM (speed) suggested that AP placement took longer than AL placement, with a mean difference of approximately 3.57 seconds. Although this was statistically significant in the small paired sample, the finding should be interpreted cautiously. The clinical importance of a delay of this magnitude is uncertain. Observational data have suggests that AP placement may be associated with improved ROSC in shockable out-of-hospital cardiac arrest, although not improved survival to admission or discharge (Lupton et al., 2024). Related cardioversion and pacing evidence has also suggested potential electrical advantages of AP positioning (Moayedi et al., 2022). However, these data do not establish whether AP placement improves meaningful outcomes during cardiac arrest.

The 2025 European Resuscitation Council guidelines recommend considering AP pad placement in refractory VF, reflecting continued uncertainty about optimal pad placement in this context (Soar et al. 2025). Recent systematic review evidence also highlights the limited and heterogeneous evidence base relating to pad size, orientation and placement during defibrillation (Ristagno et al., 2025). The present findings therefore support further research before AP placement is adopted more widely in prehospital practice.

### Strengths

A key strength of RESTART-SIM is its direct clinical relevance. The studies addressed two practical questions central to real-world resuscitation: whether AP pads can be placed accurately and whether AP positioning delays defibrillation. The staged design also allowed progression from exploratory accuracy testing to a controlled comparison of application speed.

Simulation enabled standardised assessment without patient risk. RESTART-SIM (accuracy) used anonymised identifiers and independent review of pad placement photographs, while RESTART-SIM (speed) used a randomised crossover design so that participants acted as their own controls. The use of EMS participants, standard ambulance defibrillation pads and a clothed 80-kg manikin improved the practical relevance of the speed study.

### Limitations

The principal limitation is the small and convenience sample size, meaning RESTART-SIM should be interpreted as feasibility and hypothesis-generating work rather than a definitive comparison of AL and AP pad placement. Both studies were simulation-based and therefore did not fully replicate real OHCA conditions, such as confined spaces, active CPR, patient body habitus, environmental constraints or competing clinical priorities.

RESTART-SIM (accuracy) used an unclothed manikin and fixed task order introducing several confounders. Fatigue from the attempts and the increased familiarity with the pads and manikin. As Al always preceded AP this introduced practice and repetition of placing pads along with the guided attempts preceding this. Consequently, the improvement in AL placement cannot confidently be attributed solely to the written instructions.

RESTART-SIM (speed) was conducted in a lecture-room setting where observational learning may have occurred. Age and sex were not recorded, and AL placement may have been faster because it is more familiar in routine practice.

## Conclusion

RESTART-SIM suggests that AP defibrillator pad placement is feasible in a simulated OHCA setting, but may be slower and less accurately performed than standard AL placement. Written guidance improved AL pad placement, but did not produce a similar improvement in AP accuracy, where inferior or abdominal placement remained common. Given the small sample size and simulation-based design, these findings should be interpreted as preliminary. Further work is required to refine AP-specific training, assess performance in more realistic resuscitation settings, and determine whether any potential clinical benefit of AP placement outweighs the operational delay and complexity associated with its use.

## Data Availability

All data produced in the present study are available upon reasonable request to the authors

## Appendix

### Simulation-Based Research Extensions for the STROBE Statement

**Table 4.** RESTART V1.0 Summarized categories.

| Category | A | B | C | D |
| --- | --- | --- | --- | --- |
| Pads too low / inferior / abdominal | 2 | 6 | 1 | 9 |
| Wrong side / laterality error | 1 | 1 | 0 | 1 |
| Straddling midline | 0 | 4 | 0 | 0 |
| Overlapping clavicle / too high | 4 | 0 | 0 | 0 |
| Lateral positioning issue (too medial / not lateral enough / too lateral) | 2 | 3 | 0 | 1 |
| Both pads anterior | 1 | 0 | 0 | 0 |
| Acceptable / borderline acceptable | 1 | 2 | 2 | 1 |
| Mixed error (multiple issues in same comment) | 0 | 3 | 1 | 3 |

**From: [Reporting guidelines for health care simulation research: extensions to the CONSORT and STROBE statements](https://link.springer.com/article/10.1186/s41077-016-0025-y)**

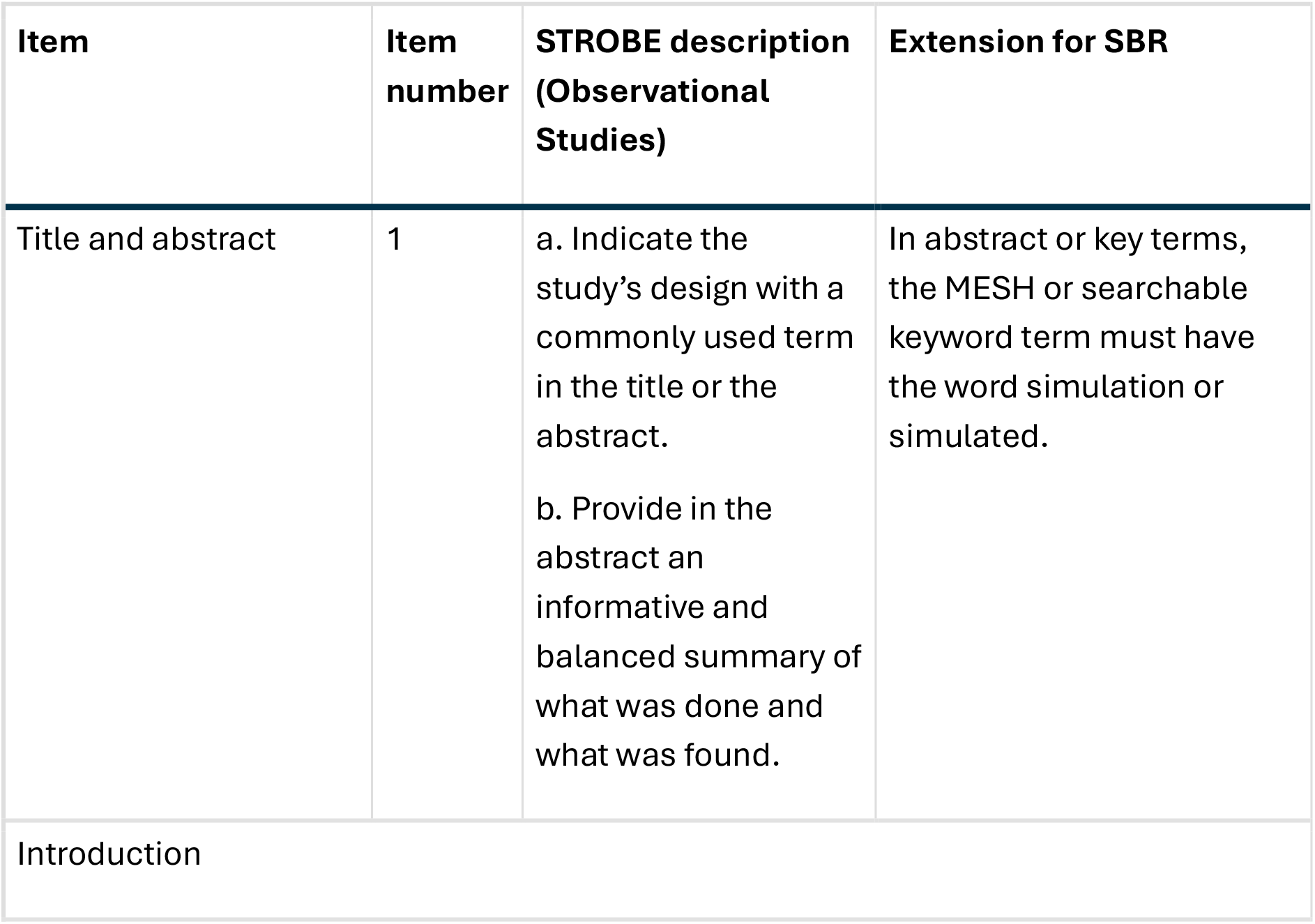
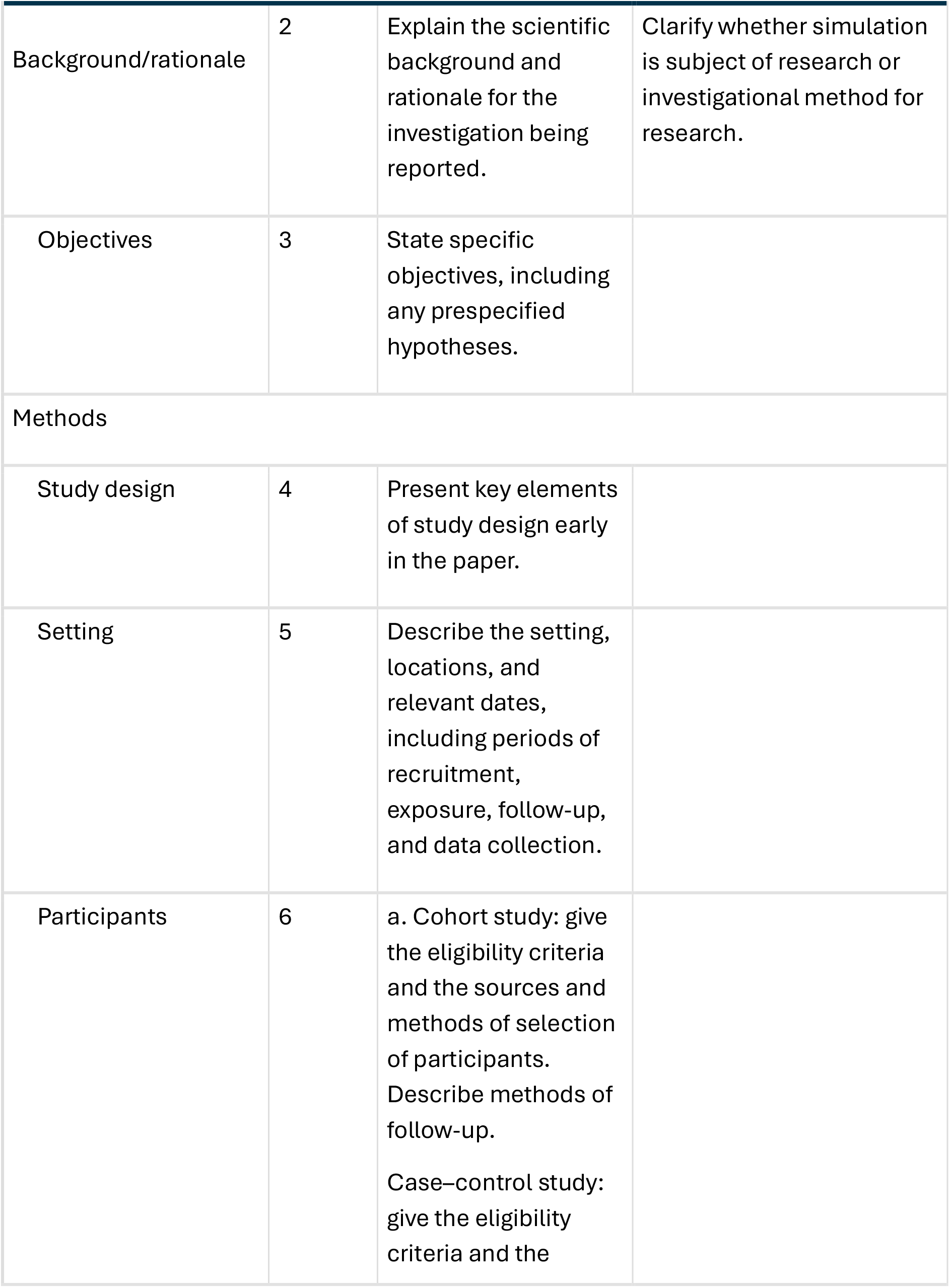
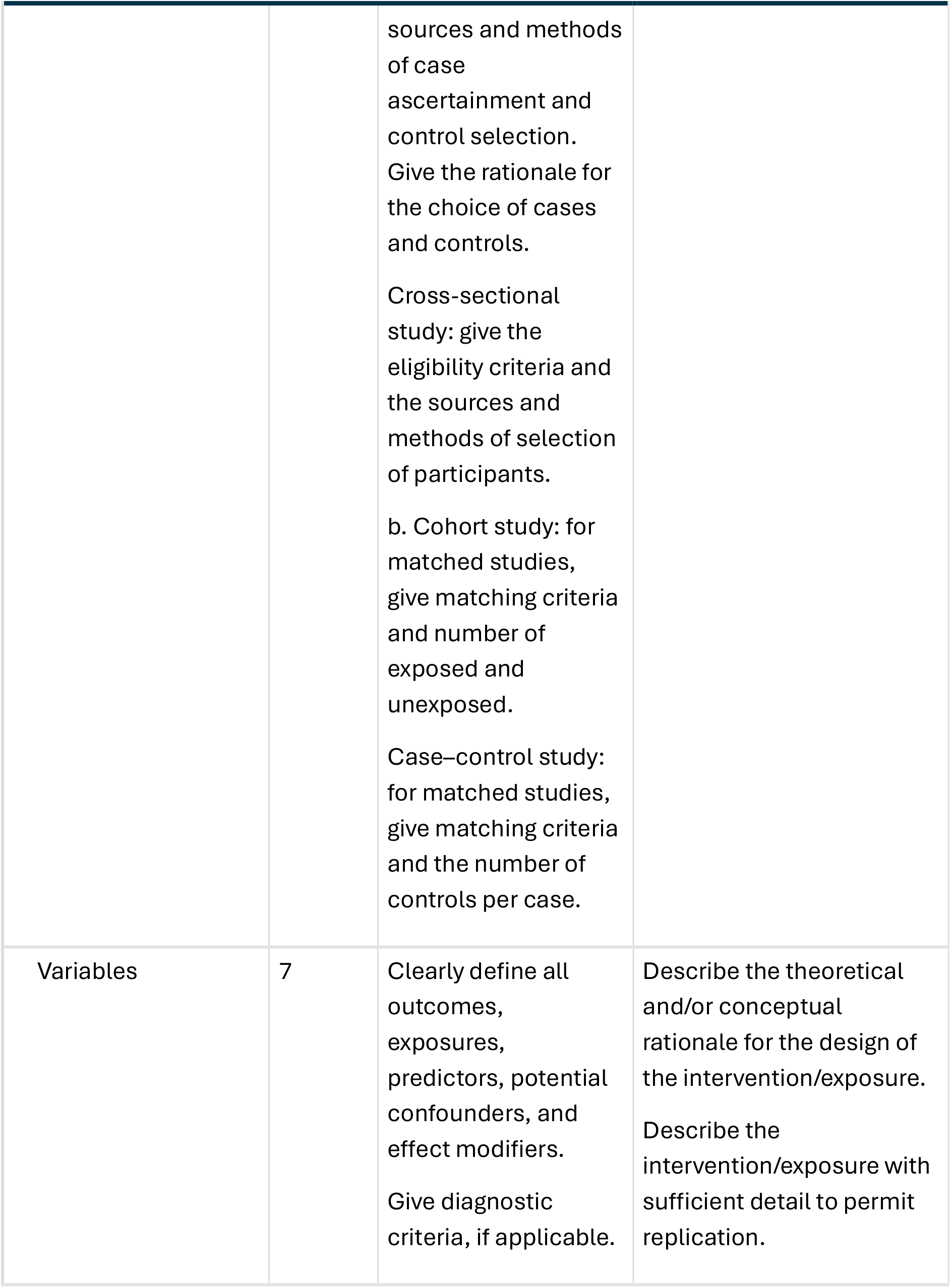
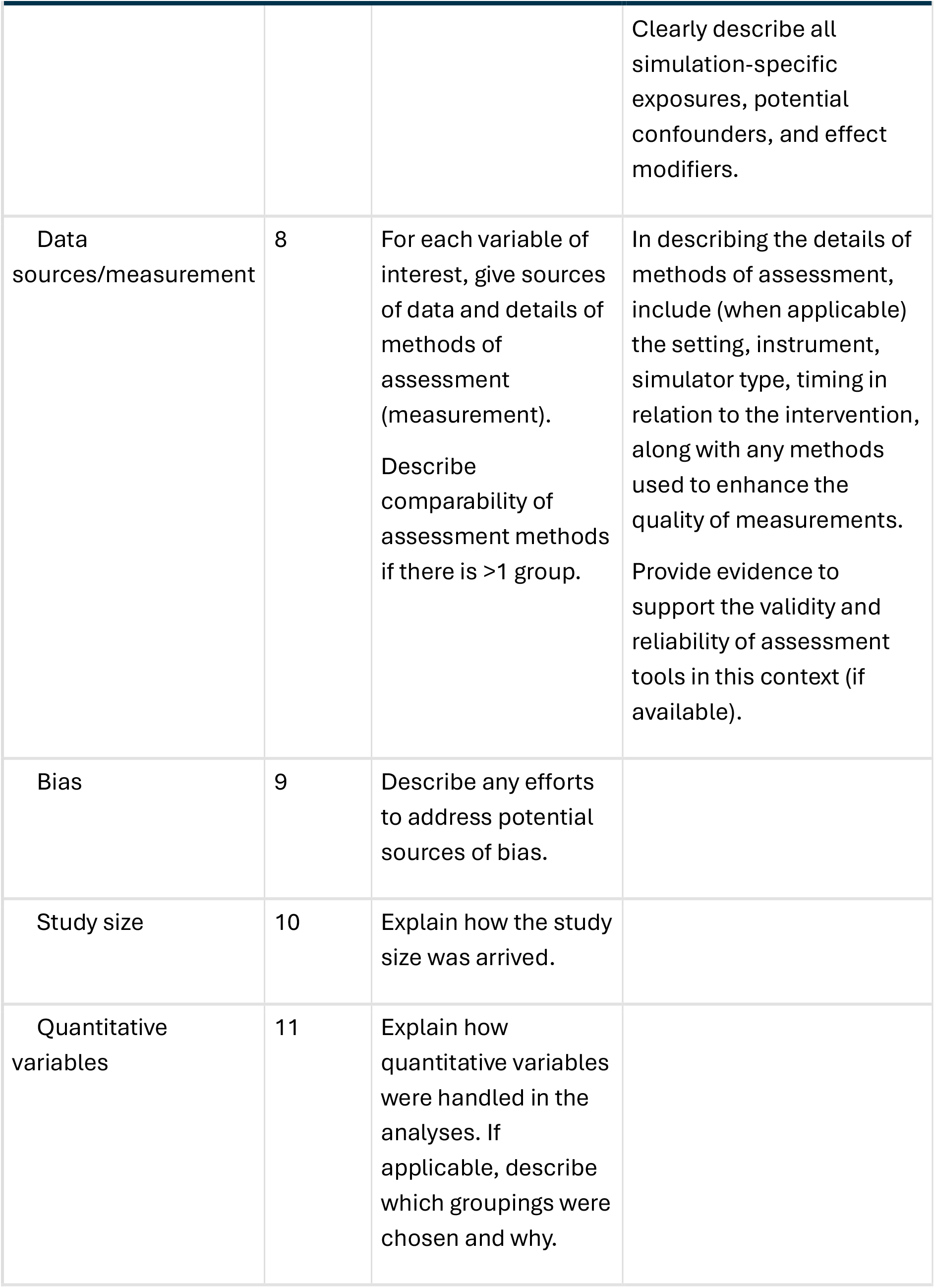
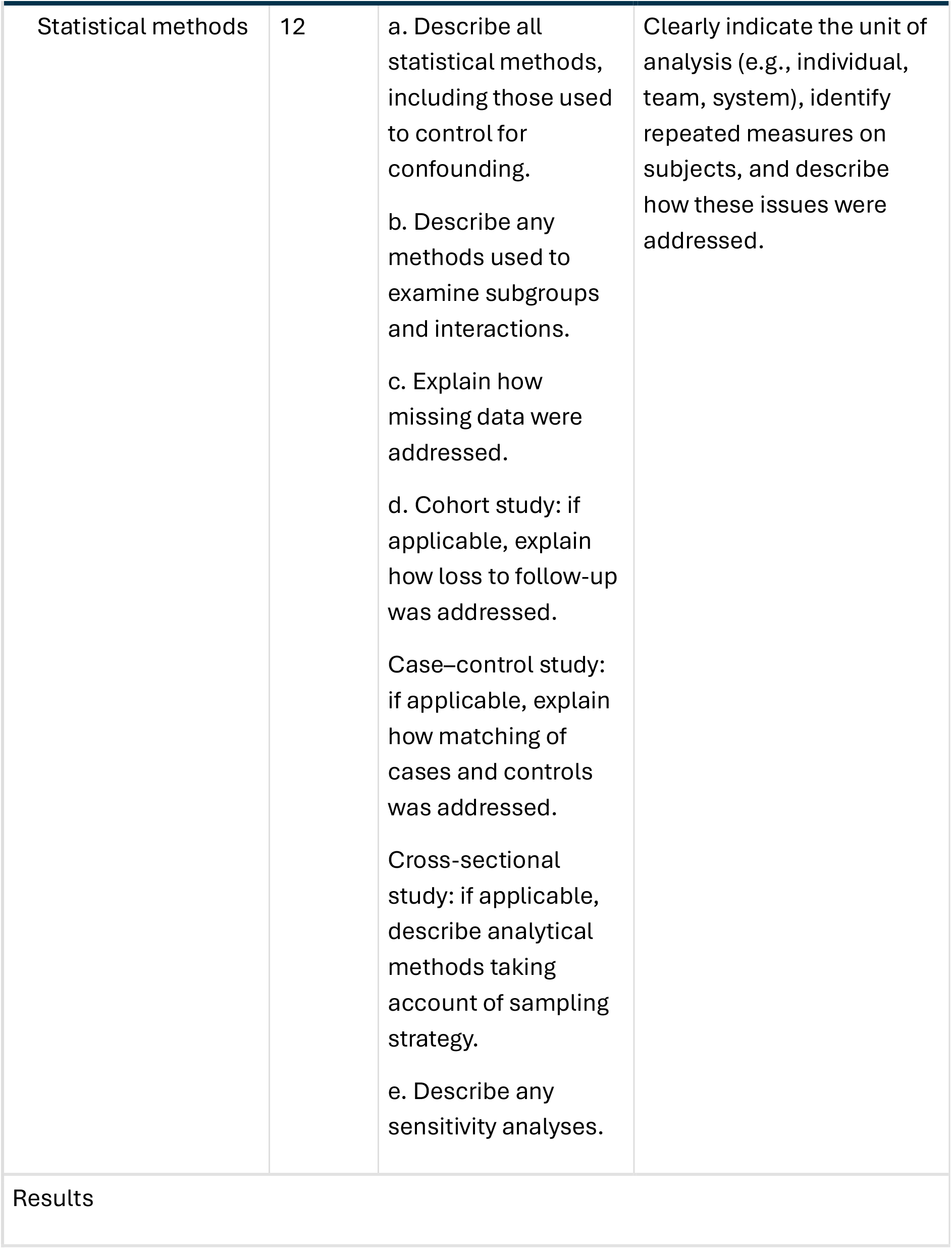
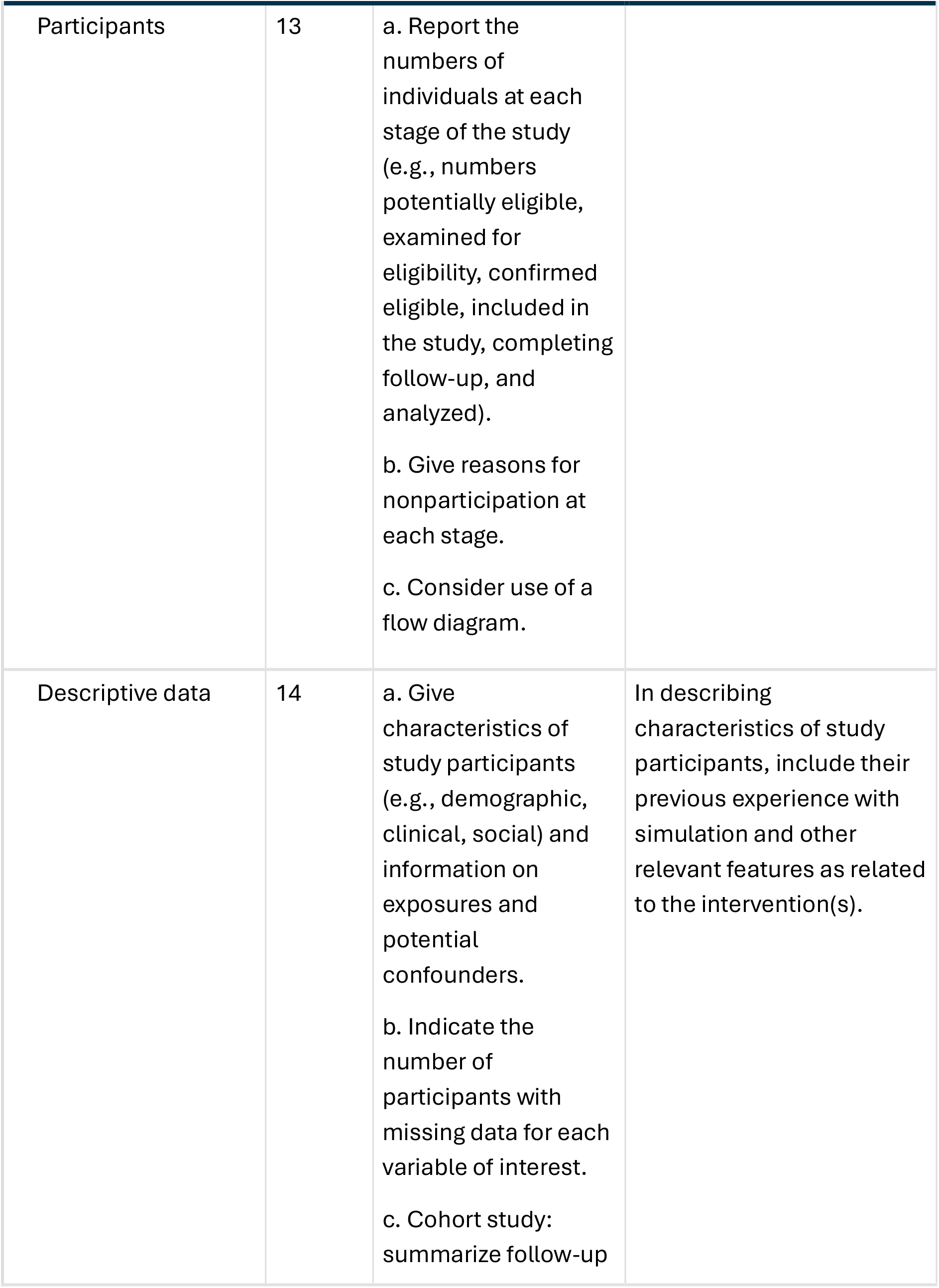
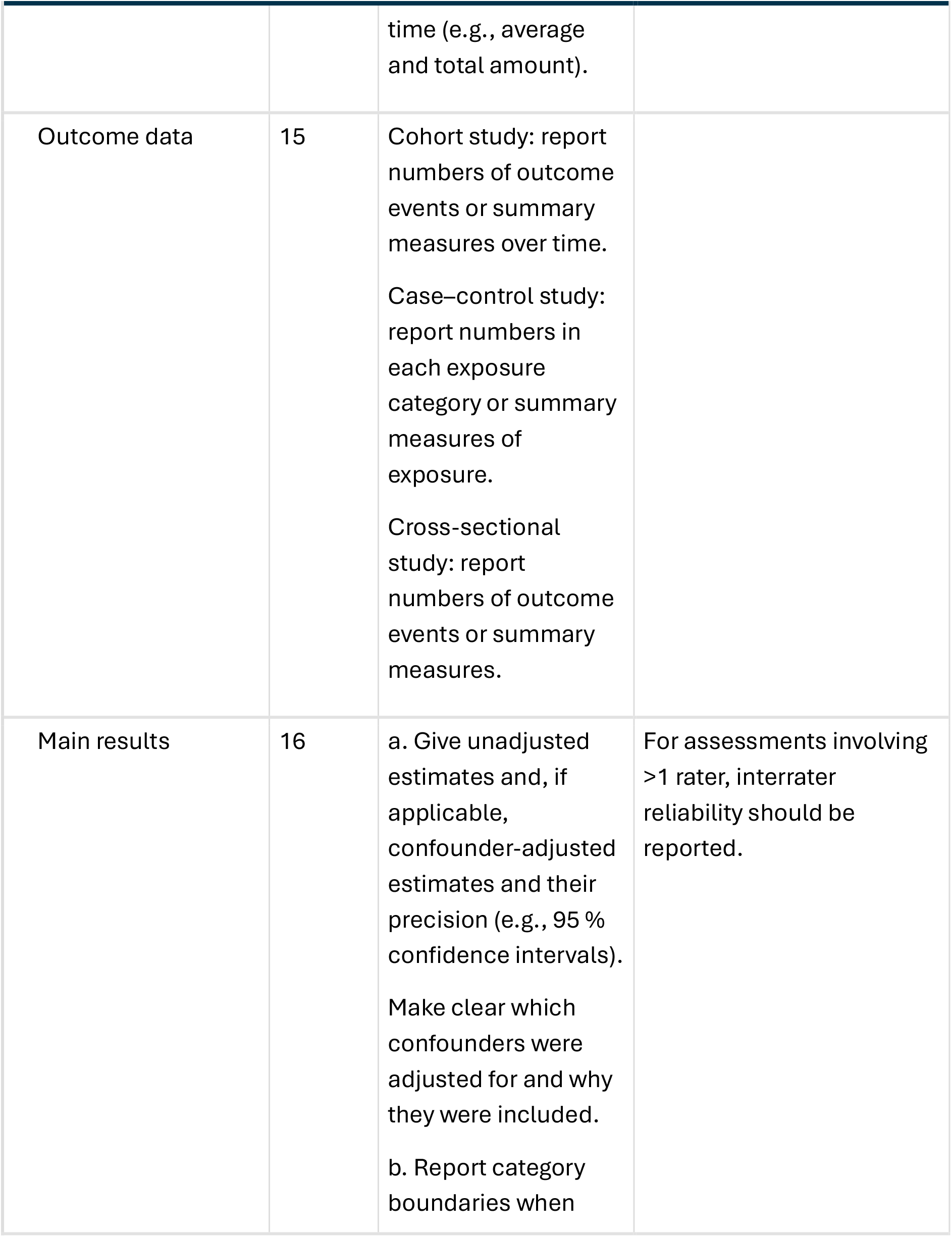
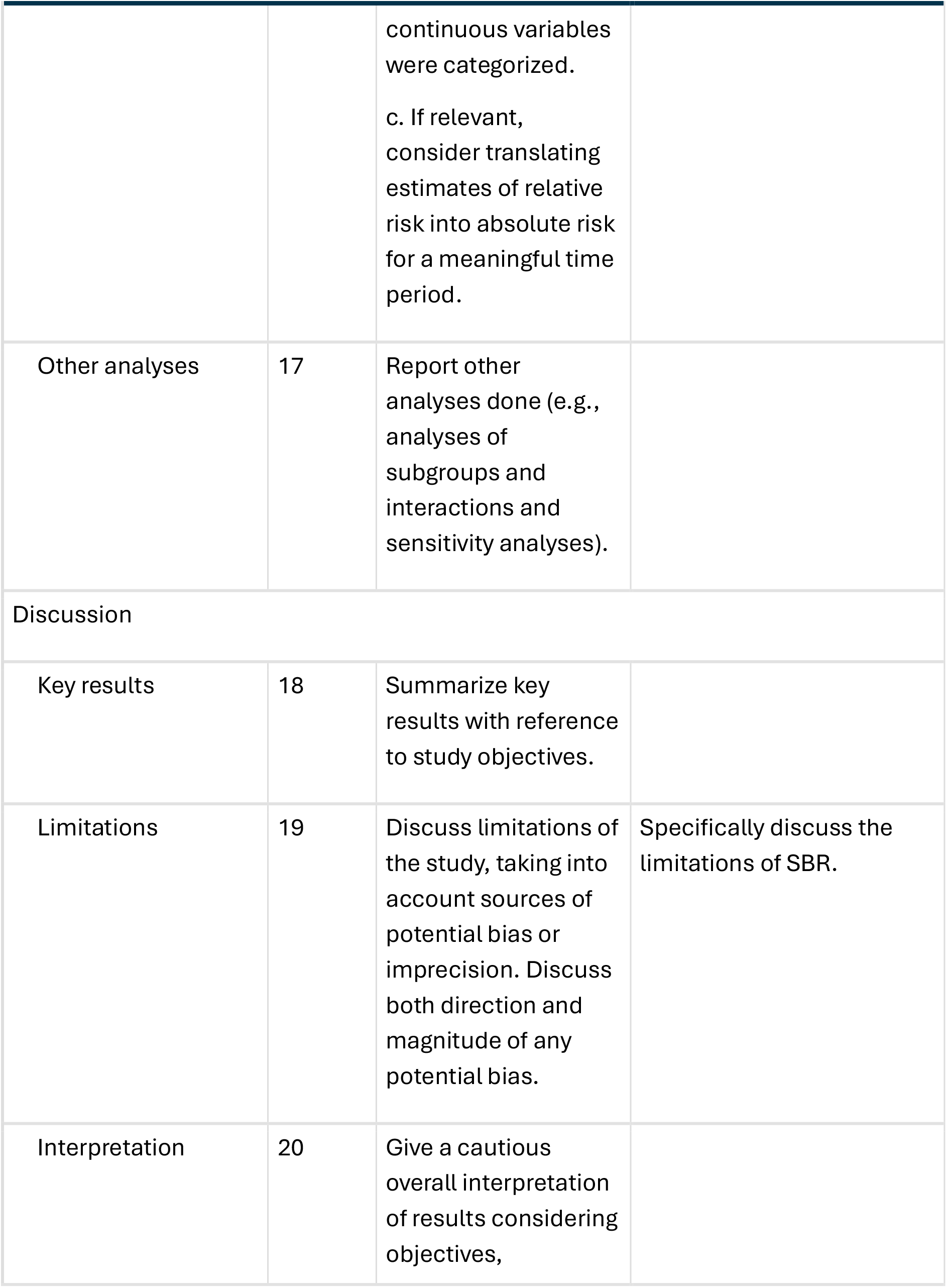
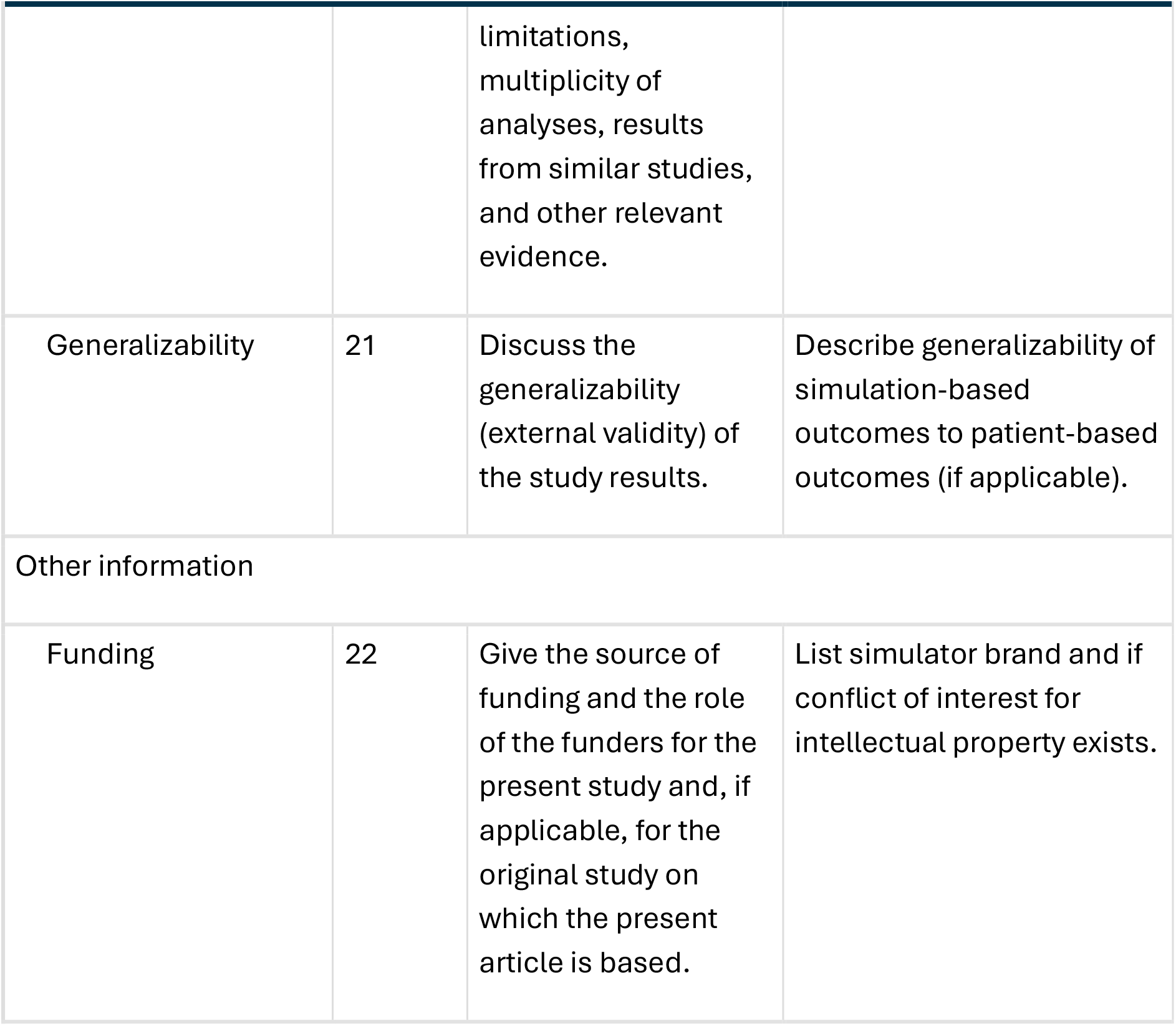

### Antero-lateral pad placement (attempt C)

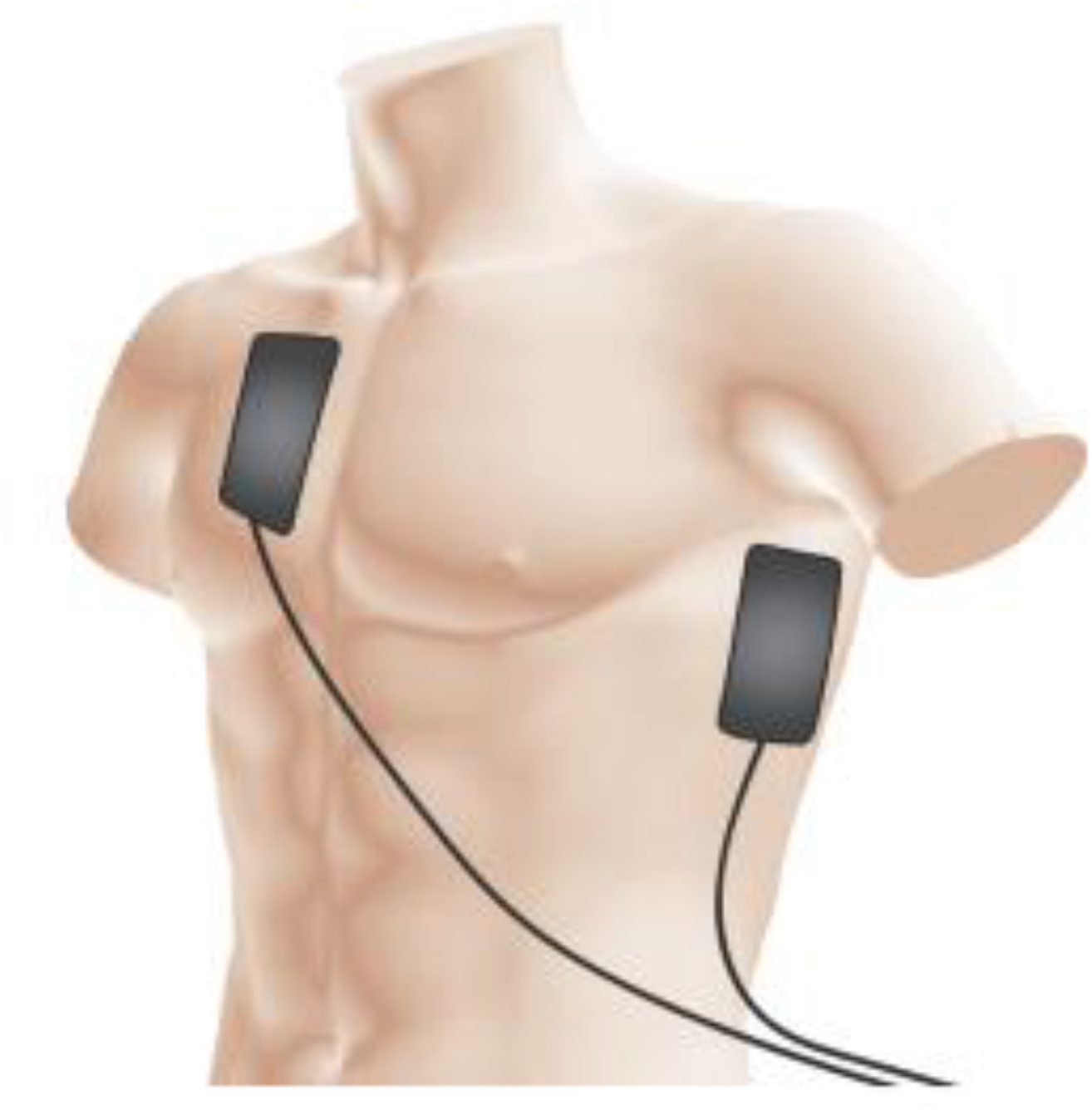

Ensure that the apical (lateral) pad is positioned correctly (mid-axillary line, level with the V6 ECG electrode position) i.e. below the armpit in the mid-axillary line

### Anterior-posterior pad placement (attempt D)

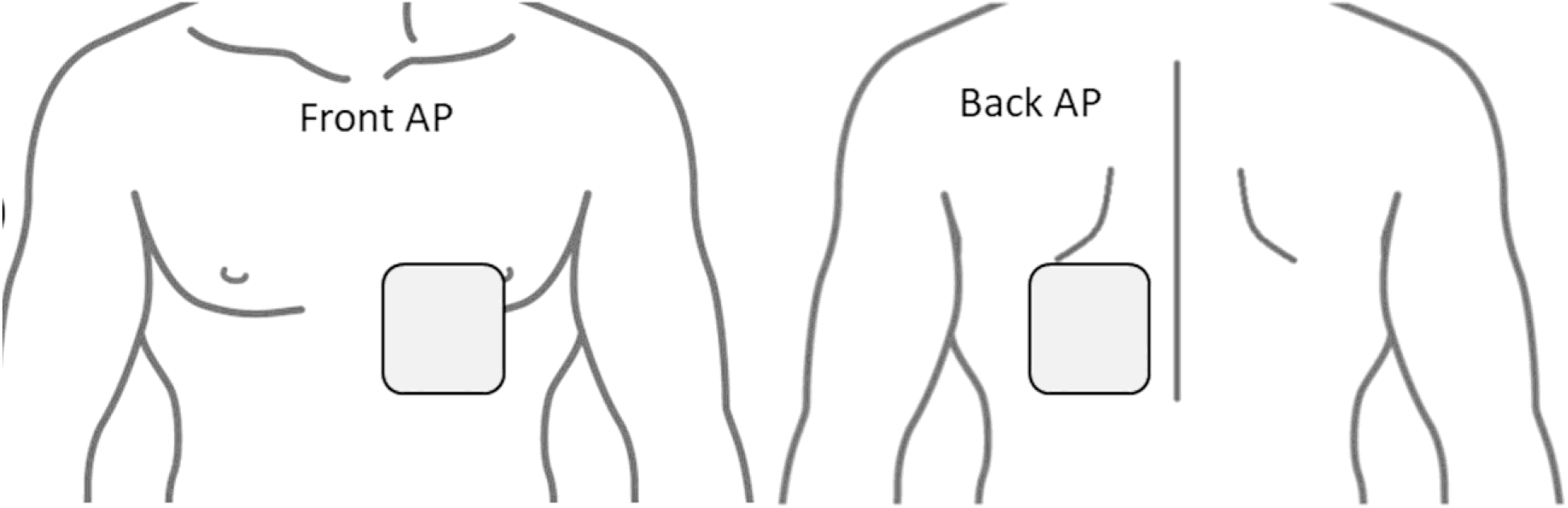

The anterior pad is placed to the left of the sternum, avoiding as much breast tissue as possible. The posterior pad is placed at the same height, centred just medial to the left scapula

